# Seroincidence of Enteric Fever in Juba, South Sudan

**DOI:** 10.1101/2022.02.28.22271422

**Authors:** Kristen Aiemjoy, John Rumunu, Juma John Hassen, Kirsten E. Wiens, Denise Garrett, Polina Kamenskaya, Jason B. Harris, Andrew S. Azman, Peter Teunis, Jessica Seidman, Joseph F. Wamala, Jason R. Andrews, Richelle C. Charles

## Abstract

We apply a new serosurveillance tool to estimate typhoidal Salmonella burden from dried blood spots in Juba, South Sudan, finding a seroincidence rate of 35 per 100 person-years and cumulative incidence of 78% over four years.

## INTRODUCTION

Enteric fever, a systemic bacterial infection caused by *Salmonella enterica* serovars Typhi and Paratyphi, causes substantial illness and mortality globally (1). However, estimating the population-level burden of infection is challenging. The reference standard for both diagnosis and surveillance, blood culture, requires microbiological laboratory facilities that are not available in many low-and-middle income countries.

Juba, the capital city of South Sudan, experiences a high burden of enteric infections such as cholera and hepatitis E virus (2,3). While enteric fever is a frequently diagnosed etiology of acute fever, there are no laboratories with blood culture capacity to confirm the diagnosis. Similarly, with no blood culture facilities, the population-level burden of enteric fever is unknown.

Hemolysin E (HlyE), a pore-forming toxin, has proven to be an accurate serologic marker for diagnosing typhoidal salmonella (4–9). New serologic tools have recently been developed to measure population-level enteric fever incidence from cross-sectional serosurveys using anti-IgG and IgA responses HlyE (10). Here, we apply these tools to generate population-level enteric fever incidence estimates in Juba, South Sudan, using banked dried blood spots collected for a SARS-CoV-2 serosurvey (11).

## METHODS

This study utilizes dried blood spots (DBS) collected from participants of a representative SARS-CoV-2 serosurvey in Juba, South Sudan enrolled between August 7^th^ and September 20^nd^, 2020 (11). Two-stage cluster sampling was used to randomly select households from predefined enumeration units within six administrative divisions within and surrounding Juba. All individuals ≥1 year of age and residing for ≥ 1 week within the sampled household were eligible to participate. Capillary blood was collected onto Whatman 903 Protein Saver cards, air dried and transported at ambient temperature to Massachusetts General Hospital where they were stored at 4°C. We tested all banked samples collected from participants under age 25 and a random sample of participants 25 and older. The study protocol was approved by ethical review boards with the South Sudan Ministry of Health and Mass General Hospital.

We used kinetic enzyme-linked immunosorbent assays to quantify antibody levels present in eluted DBS as previously described (5,6). We submerged two 6 mm2 punches from the DBS in 133 *μ*L of PBS-0.05% Tween 20r overnight at 4°C with gentle agitation, and recovered eluate after centrifugation. The eluate was stored frozen at −20 C until testing.

We used the antibody dynamics modeled from a longitudinal cohort of 1410 blood culture-confirmed enteric fever cases (10) to estimate enteric fever incidence as described in (12–14). In brief, we created a likelihood function for the observed cross-sectional population data based on antibody dynamics following blood-culture confirmed infection. We generated joint incidence estimates by combining the likelihood for HlyE IgA and IgG for each age stratum (0-4yr, 5 - 15yr, ≥16yr) using age-specific antibody dynamics. This method incorporates heterogeneity in antibody responses and explicitly accounts for measurement error and biologic noise as detailed in (10). We used the same method to generate individual-level incidence estimates based on quantitative HlyE IgA and IgG responses and used the exponential probability distribution to calculate the two-year and four-year cumulative incidence. We then fit age-dependent curves to using generalized additive models (15) with a cubic spline for age and simultaneous confidence intervals using a parametric bootstrap of the variance-covariance matrix of the fitted model parameters (16). We compared our results to the seroprevalence derived from a mean plus three standard deviation cutoff using North American control population of 1) 48 children aged 1-5 years who had first degree relatives with celiac disease, enrolled nationally and 2) 31 healthy controls, children and young adults aged 2-18 years, enrolled at Massachusetts General Hospital (17). Code and de-identified data are available on (<u>link_ToBeAdded_github_repo</u>).

## RESULTS

2214 individuals were enrolled and provided blood samples, 1840 had complete interview data and 1290 were randomly selected for testing (11). The median age of tested participants was 17 years old (interquartile range (IQR) 10-24), and 63.5% (819/1290) were female (Table 1).

**Table 1:** Summary of population.

|  | Overall<br>(N=1290) |
| --- | --- |
| **Sex** |  |
| Female | 819 (63.5%) |
| Male | 471 (36.5%) |
| **Age, in years** |  |
| Median (IQR) | 17 (10-24) |
| **Age category, in years** |  |
| 1-3 | 41 (3.2%) |
| 4-6 | 118 (9.1%) |
| 7-9 | 134 (10.4%) |
| 10-14 | 259 (20.1%) |
| 15-24 | 423 (32.8%) |
| 25-34 | 167 (12.9%) |
| 35-44 | 62 (4.8%) |
| 45+ | 86 (6.7%) |

**Table 2:** Age-dependent incidence rates and cumulative incidence.

| Age, in years | Incidence rate per 100 person-years (95% CI) | Two-year Cumulative Incidence (IQR) | Four-year Cumulative Incidence (IQR) |
| --- | --- | --- | --- |
| 1-3 | 51.6 (46.3-70.7) | 57.9% (50.1-78.4) | 82.2% (75.1-95.4) |
| 4-6 | 35.2 (32.5-44.3) | 59.1% (37.0-70.1) | 83.3% (60.3-91.0) |
| 7-9 | 33.2 (31.0-40.5) | 52.5% (42.6-62.0) | 77.4% (67.0-85.6) |
| 10-14 | 27.8 (26.4-32.4) | 48.1% (34.6-61.6) | 73.1% (57.2-85.2) |
| 15-24 | 31.6 (27.5-47.2) | 47.5% (33.6-65.3) | 72.5% (55.9-87.9) |
| 25-34 | 33.7 (31.3-42.1) | 56.3% (40.2-69.8) | 80.9% (64.2-90.9) |
| 35-44 | 45.8 (40.4-65.9) | 61.3% (54.6-71.9) | 85.0% (79.4-92.1) |
| 45+ | 39.0 (35.1-53.0) | 56.5% (45.1-72.2) | 81.1% (69.9-92.3) |
| Overall | 35.0 (32.4-37.9) | 52.7% (37.2-67.3) | 77.6% (60.6-89.3) |

Median HlyE IgG (10.4, IQR 6.1 – 12.7) and IgA (3.5, IQR 2.3 – 5.2) responses were elevated well above a North American pediatric control population (IgG: 0.16, IQR 0.07 – 0.35; IgA: 0.3, IQR 0.001 – 0.92) and were comparable to responses observed among blood-culture confirmed enteric fever cases 8-12 months after symptom onset (IgG: 12, IQR 5.9– 24; IgA: 4.4, IQR 2.2 – 9.4) (10) (Figure 1).

**Figure 1:**
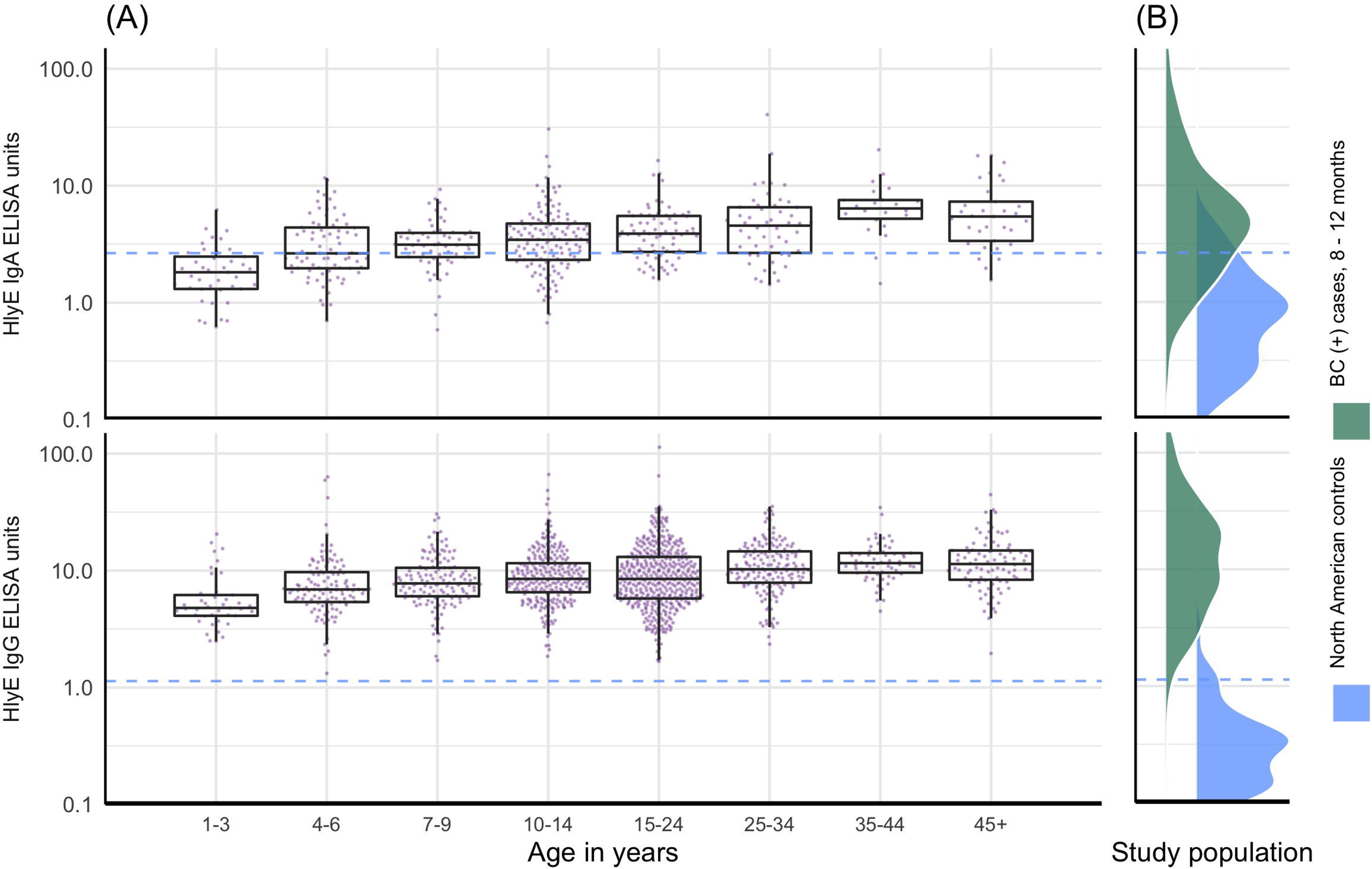
Age-dependent anti-hemolysin E (HlyE) IgA (top) and IgG (bottom) responses in Juba population sample compared to blood culture confirmed cases between 8 and 12 months from fever onset and North American controls. (A) Cross-sectional antibody responses to HlyE IgA and IgG according to age in years measured from a serosurvey of 1290 individuals in Juba, South Sudan collected between August 7^th^ and September 2^nd^, 2020. Each point is an individual sample. (B) Density of antibody responses among 1410 blood-culture confirmed enteric fever cases in Bangladesh, Nepal, Pakistan and Ghana between 8 and 12 months after symptom onset as reported in (10) and anegative control population of 1) 48 children aged 1-5 years who had first degree relatives with celiac disease, enrolled nationally and 2) 31 healthy controls, children and young adults aged 2-18 years, enrolled at Massachusetts General Hospital (17). The dashed blue line across all three panels represents the mean plus three standard deviations of HlyE IgA and IgG values observed in the pediatric control population.

The age-specific enteric fever incidence estimates per 100 person-years ranged from 27.8 (95% confidence interval (CI): 26.4 – 32.4) among children 10 to 14 years old to 51.6 (95% CI: 46. – 70.7 among children 1 to 3 years old (Figure 2). The overall incidence rate was 35.0 (95% CI 32.4-37.9). The cumulative incidence over two years was 52.7% (IQR 37.2-67.2) and over 4 years was 77.6% (IQR 60.6 - 49.3). Using a cutoff derived from a North American pediatric control population, 99.8% (1288/1290) of the population were seropositive using HlyE IgG and 65.2% (318/488) were positive using HlyE IgA (Supplemental Figure 1).

**Figure 2:**
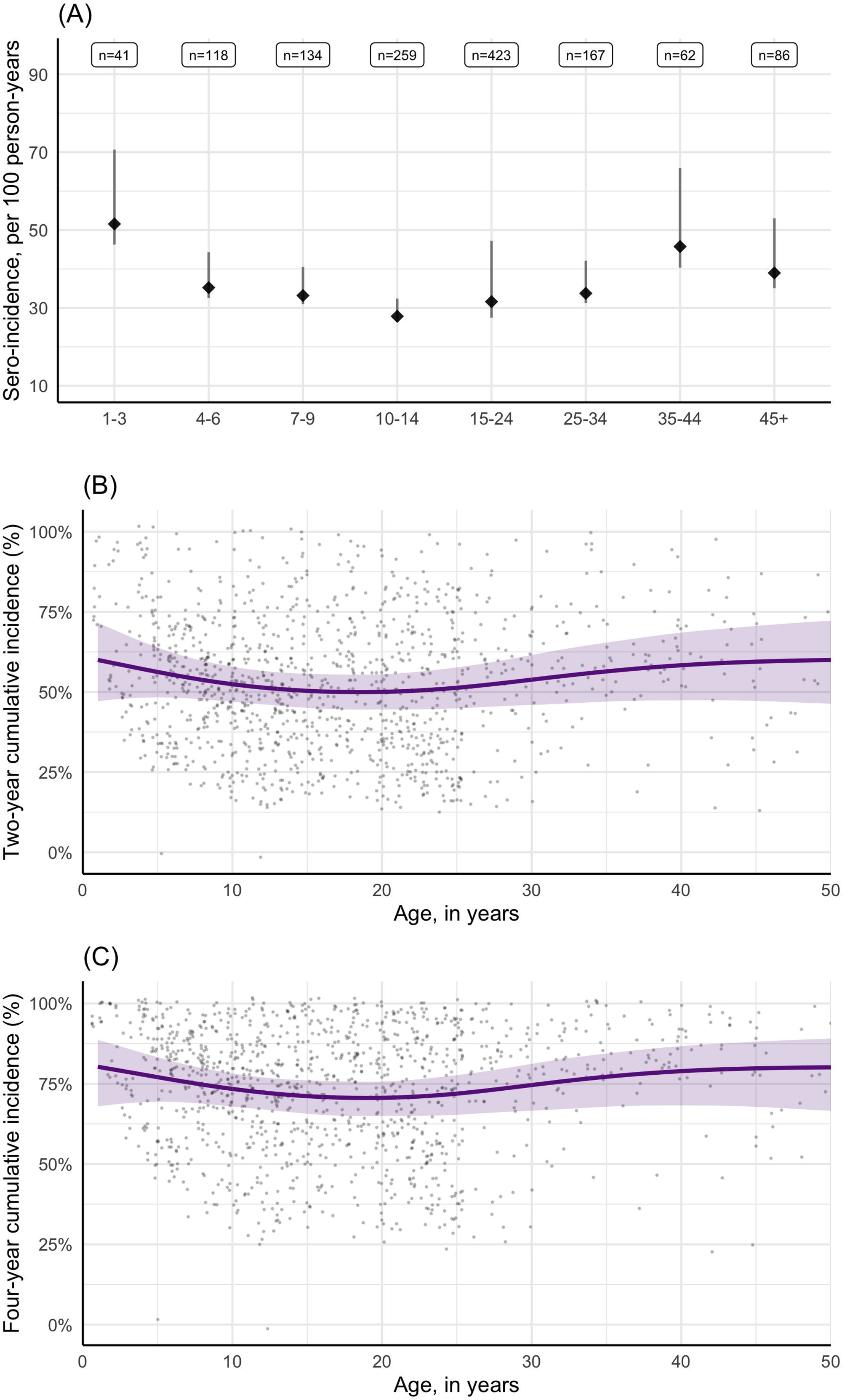
Estimated sero-incidence of typhoidal Salmonella by age group in Juba, South Sudan. (A) Sero-incidence per age-group with 95% credible intervals. (B & C) Individually-predicted incidence estimates (points) and the smoothed cumulative incidence over two years (B) and four-years (C).

## DISCUSSION

Here we apply a new serosurveillance tool to estimate the population-level incidence of enteric fever in Juba, South Sudan. Using banked DBS collected for a SARS-CoV-2 serosurvey, we rapidly estimated the burden of typhoidal S*almonella* in a region with no blood culture surveillance and no population-level enteric fever incidence estimates. We estimated a high incidence rate of 35.0 infections per 100 person-years, with over 75% of the sampled population infected in the prior 4 years.

The analytic approach is an improvement over cutoff based methods in that we can combine information from both HlyE IgA and IgG responses to generate a consensus incidence estimate, while accounting for heterogeneity in antibody responses, measurement error and biologic noise. While the cutoff-based method yielded a seroprevalence of nearly 100% for HlyE IgG, we generated cumulative incidence estimates over a precise time window and could identify populations with recent and later infections.

There are limitations to this study which should be considered when interpreting the results. First, only 1840 samples of the 2214 enrolled had linked age data. Second, internally-displaced person camps were not included in the serosurvey. As displaced persons have been identified as high-risk populations for enteric infections, it would be valuable to include them in future enteric fever surveillance studies to determine if this population is at higher or equivalent risk (2). Finally, we used longitudinal antibody kinetics estimates from enteric fever cases in Bangladesh, Pakistan, Nepal and Ghana to estimate incidence in South Sudan. While we did not observe overt differences in the kinetics of antibody responses across countries (10), it may still be possible that the decay rate among enteric fever cases in Juba is different because of the high force of infection and differences in exposure to other infections.

Our results suggest a high burden of enteric fever in Juba, South Sudan warranting urgent public health and research attention. The seroincidence tool we use can be applied to other regions lacking blood culture surveillance to generate rapid enteric fever incidence estimates, providing the high-resolution data critically needed to inform typhoid conjugate vaccine introduction.

## Data Availability

All data produced in the present study are available upon reasonable request to the authors

## FIGURE LEGENDS

**Supplemental Figure 1:**
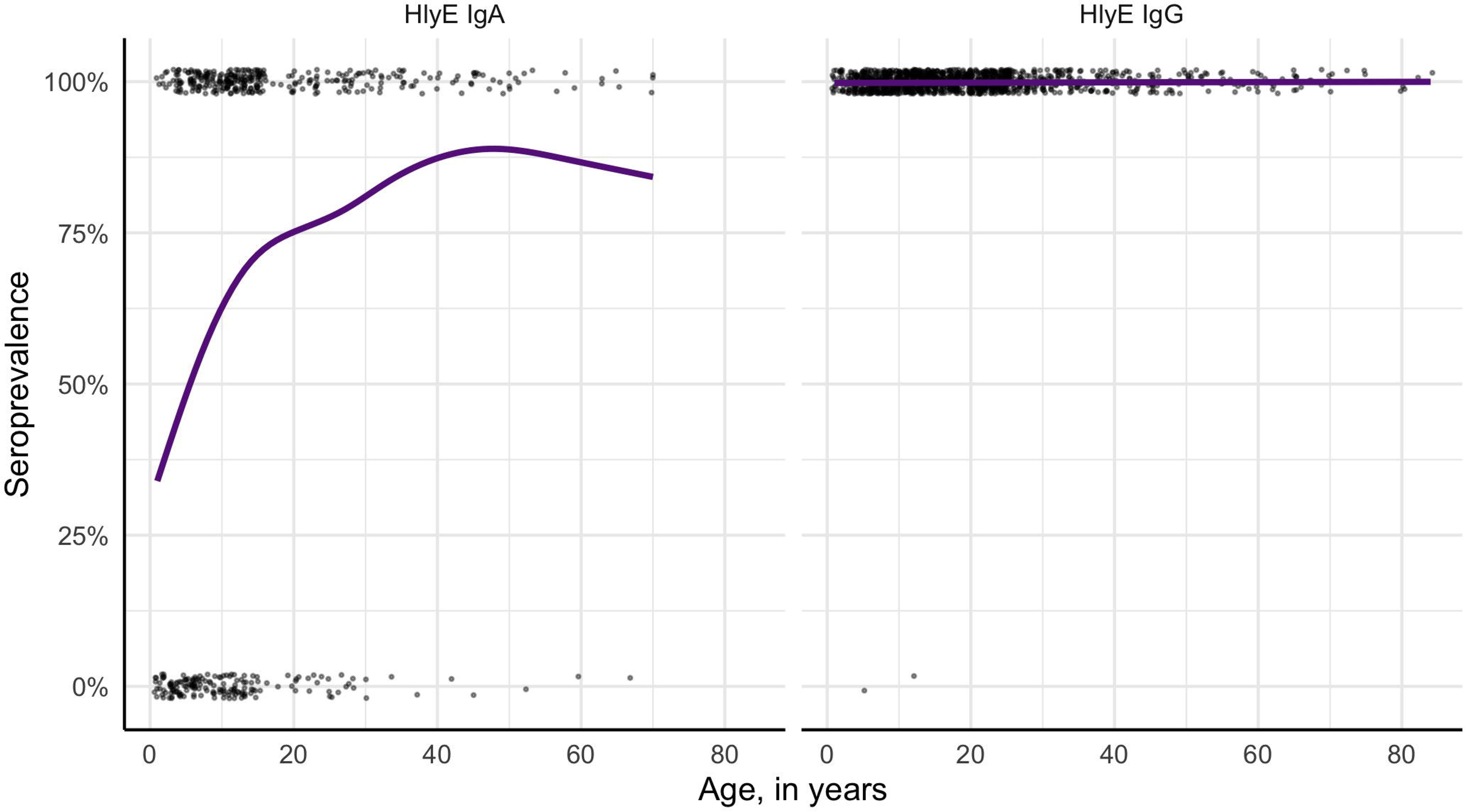
Age dependent seroprevalence curves using negative control cutoff. Each point is an individual classified into positive of negative based on a cutoff derived from a North American control population for HlyE IgA (left) and HlyE IgG (right). The purple line is the age-dependent sero-prevalence curve fit using generalized additive models.

